# Relationship between COVID-19 pandemic and ecological, economic, and social characteristics

**DOI:** 10.1101/2021.03.22.21254134

**Authors:** Attila Murányi, Bálint Varga

**Author notes:** Corresponding author. Attila Murányi, D.Sc.

## Abstract

COVID-19 pandemic had huge impacts on the global world, with both a negative impact on society and economy, but a positive one on nature. But this universal effect resulted in different infection rates from country to country. We analyzed the relationship between the pandemic and ecological, economic, and social characteristics. All of these data were collected in 140 countries at 6 time points. Correlations were studied using univariate and multivariate regression models.

The world was interpreted as a single global ecosystem consisting of ecosystem units representing countries. We first studied 140 countries around the world together, and infection rates were related to per capita GDP, Ecological Footprint, median age, urban population, and Biological Capacity, globally. We then ranked 140 countries by infection rate and created 4 equal groups, each with 35 countries. In the first group, the infection rate was very high and was related to the Ecological Footprint (consumption) and GDP per capita (production). This group is dominated by developed countries and their ecological characteristics have proven to be particularly significant. In groups 2, 3, and 4, infection rates were high, moderate, and low, and were primarily associated with median age and urban population.

In the scientific discussion, we have interpreted why infection is high in developed countries. Sustainable ecosystems are balanced, unlike the ecosystems of developed countries. According to science, the resilience and health of both natural ecosystems and humans are closely linked to the world of microbial communities. Our results suggest that both the economy and society need to be in harmony with nature, creating sustainable ecosystems in developed countries as well.

## Introduction

The first wave of the COVID-19 pandemic shocked humanity and severely limited the functioning of society and the economy. Many scientists believe that the COVID-19 pandemic is a stage in a process, a logical consequence of the degradation and depletion of nature. Can the COVID-19 pandemic be interpreted as a reaction of nature? We hypothesize that the characteristics of the COVID-19 pandemic are not independent of ecological, economic and social conditions.

For the global analysis of a pandemic, we interpreted the world as a single global ecosystem consisting of ecosystem units representing countries. The natural and environmental condition of the country can be described by ecological characteristics. However, a country can be described not only by ecological but also by economic and social characteristics.

The COVID-19 coronavirus is a biological agent from nature and has had a major impact on the global world (i.e., the global ecosystem) as well as individual countries (i.e., individual ecosystem units). In principle, the rate of COVID-19 coronavirus infection should be the same in all countries, but this is not the case. We hypothesized that the effect of COVID-19 in different countries would depend not only on the virus and its spread, but also on ecological, economic, and social conditions. Although every country is made up of nature, the economy and human society, nature is under pressure from our ever-increasing production and ever-increasing consumption.

## Materials and methods

The pandemic was analyzed during the first wave. Pandemic data were downloaded from the https://www.worldometers.info/coronavirus/ website at 6 time points (April 18, May 2, May 16, May 30, June 18, July 4). The Worldometer provides live statistics and official real-time data on the COVID-19 pandemic from all over the world [1]. The use of relative data to compare different countries was preferred, so the effect of COVID-19 coronavirus was characterized by the infection rate. The infection rate is the total number of cases per million people reported by countries.

The state of nature and the state of the environment were characterized by the ecological conditions of the country. Ecological conditions were characterized by Ecological Footprint and Biological Capacity. The Ecological Footprint per person is defined as the area used to support the consumption of the country divided by the population. Biological Capacity of a given country is the ecosystem’s capacity to produce biological materials used by the country and to absorb waste material generated by the country. Biological Capacity per person is the Biological Capacity of the country divided by the population. Their unit of measurement is the global hectares per person [2].

The economy was characterized by GDP per capita. GDP per capita is gross domestic product divided by midyear population. GDP is the sum of gross value added by all resident producers in the economy plus any product taxes and minus any subsidies not included in the value of the products. It is calculated without making deductions for depreciation of fabricated assets or for depletion and degradation of natural resources. Data are in current U.S. dollars [3]. Society was characterized by the median age of the population (year), urban population (%), population density (persons per square km), number of inhabitants and migrants [4]. To study the global ecosystem, we were able to use data from a total of 140 countries in which all of these data were available.

The correlations between the infection rate and the ecological, economic, social characteristics were studied by univariate and multivariate regression models. In case of multiple regression, backward elimination process was used to select the most significant predictor set from the 8 initial variables, using 0.05 p-value as stopping criterion. The fit of the model was characterized by R^2^, F- and P-values from the ANOVA table. The calculations were made using the STATISTICA software.

## Results

### Infection rates on a global scale

All 140 countries were taken into account in characterizing the global ecosystem. Data from 140 countries were used at 6 time points.

We first used a univariate model to characterize the relationships between infection rate and individual ecological, economic, and social characteristics. The results were ranked according to R^2^ (Table 1). The infection rate can best be explained by GDP per capita, Ecological Footprint, median age, and urban population during the first wave of the COVID-19 epidemic. The importance of each variable has changed over time, which is also reflected in the R2 ranking. For example, as the epidemic progressed, the importance of the population living in urban areas increased. The strongest correlation was obtained on April 18, 2020 (R^2^ = 0.7096) between infection rate and GDP/capita, when the regression equation could explain 70.96 % of the infection rate. In case of GDP/capita, Ecological Footprint, median age and urban population, R^2^ ranged from 0.30 to 0.71, 0.45 to 0.59, 0.24 to 0.36 and from 0.28 to 0.37. These results show the relationship between infection rate and ecological, economic, and social characteristics.

**Table 1.** Univariate regression equations between infection rate and ecological, economic, social characteristics on a global scale (140 countries, 6 time points).

| Regression equation | R2 | F value | P value | Regression equation | R2 | F value | P value |
| --- | --- | --- | --- | --- | --- | --- | --- |
| **18-Apr-2020** |  |  |  | **30-May-2020** |  |  |  |
| Cases/1M = 0.0328*GDP/capita | 0.7096 | 340 | 3.86E-39 | Cases/1M = 436.4*Ecofootprint | 0.5334 | 159 | 8.97E-25 |
| Cases/1M = 176.2*Ecofootprint | 0.5033 | 141 | 7.18E-23 | Cases/1M = 0.0646*GDP/capita | 0.4749 | 126 | 3.53E-21 |
| Cases/1M = 17.71*Median age | 0.3340 | 70 | 6.20E-14 | Cases/1M = 2096*Urban pop. | 0.3211 | 66 | 2.40E-13 |
| Cases/1M = 862.8*Urban pop. | 0.3151 | 64 | 4.47E-13 | Cases/1M = 39.29*Median age | 0.2839 | 55 | 1.04E-11 |
| **2-May-2020** |  |  |  | **18-Jun-2020** |  |  |  |
| Cases/1M = 0.0428*GDP/capita | 0.6958 | 318 | 9.76E-38 | Cases/1M = 593.1*Ecofootprint | 0.4589 | 118 | 2.86E-20 |
| Cases/1M = 245.2*Ecofootprint | 0.5618 | 178 | 1.11E-26 | Cases/1M = 0.0804*GDP/capita | 0.3432 | 73 | 2.33E-14 |
| Cases/1M = 24.00*Median age | 0.3535 | 76 | 7.70E-15 | Cases/1M = 2916*Urban pop. | 0.2896 | 57 | 5.88E-12 |
| Cases/1M = 1198*Urban pop. | 0.3501 | 75 | 1.11E-14 | Cases/1M = 53.08*Median age | 0.2413 | 44 | 6.22E-10 |
| **16-May-2020** |  |  |  | **4-Jul-2020** |  |  |  |
| Cases/1M = 0.0532*GDP/capita | 0.6181 | 225 | 7.59E-31 | Cases/1M = 719.4*Ecofootprint | 0.4532 | 115 | 5.96E-20 |
| Cases/1M = 331.1*Ecofootprint | 0.5884 | 199 | 1.40E-28 | Cases/1M = 3679*Urban pop. | 0.3094 | 62 | 8.03E-13 |
| Cases/1M = 1606*Urban pop. | 0.3613 | 79 | 3.26E-15 | Cases/1M = 0.0931*GDP/capita | 0.3086 | 62 | 8.71E-13 |
| Cases/1M = 31.14*Median age | 0.3417 | 72 | 2.74E-14 | Cases/1M = 66.33*Median age | 0.2529 | 47 | 2.08E-10 |

As a second step, we analyzed the global ecosystem using multivariate analysis (Table 2). The infection rate during the first wave of the COVID-19 epidemic can be explained by 5 characteristics: GDP / capita, Ecological Footprint, median age, urban population, and Biological Capacity. Four regression equations contain 2 or 3 independent variables that result in the best fit. The relationship between infection rate and ecological, economic, and social characteristics resulted in a better correlation when multivariate analysis was used. However, for 140 countries, the values of each characteristic cover a very wide range, so the application of these regression equations is not recommended for each country. Of course, these relationships are not causal, but show that the rate of infection is not independent of ecological, economic, and social conditions.

**Table 2.** Multivariate regression equations between infection rate and ecological, economic, social characteristics on a global scale (140 countries, 6 time points).

| Date | Regression equation | R2 | F value | P value |
| --- | --- | --- | --- | --- |
| 18-Apr-2020 | Cases/1M = 0.0328*GDP/capita | 0.7096 | 340 | 3.86E-39 |
| 2-May-2020 | Cases/1M = 0.0428*GDP/capita | 0.6958 | 318 | 9.76E-38 |
| 16-May-2020 | Cases/1M = 0.0297*GDP/capita + 275.8*Ecofootprint - 14.93*Median age | 0.6715 | 93 | 5.91E-33 |
| 30-May-2020 | Cases/1M = 679.1*Ecofootprint - 30.78*Median age - 27.02*Biocapacity | 0.5919 | 66 | 1.59E-26 |
| 18-Jun-2020 | Cases/1M = 918.2*Ecofootprint - 45.92*Median age | 0.5017 | 69 | 1.34E-21 |
| 4-Jul-2020 | Cases/1M = 969.5*Ecofootprint + 3152*Urban pop. - 98.58*Median age | 0.5000 | 46 | 1.61E-20 |

The correlations (R^2^) decreased as the pandemic progressed, which may be caused by a number of factors. On the one hand, we selected only eight characteristics that we considered to be the most significant. On the other hand, neither the characteristics of the spread of the epidemic nor the effects of the epidemic management were taken into account. Thus, the increasing values of (1 – R^2^) might be explained by external factors that were not taken into account.

In the global study of the COVID-19 pandemic, 140 countries form a single group, which is best characterized by the averages of each characteristic. The average for 140 countries was as follows: GDP/capita = 14,647 USD/capita; Ecological Footprint = 3.2 gha/person; Biological Capacity = 3.9 gha/person; Urban population = 60 % and Median age = 31 year. The 140 countries studied were the same, so these averages were constant at 6 time point.

### Infection rate in different country-groups

We wanted to analyze in more detail the global relationship between infection rate and ecological, economic, and social characteristics. We were wondering why the infection rate varies from country to country. We ranked 140 countries by infection rate and created 4 equal groups, each with 35 countries. In the first group of 35 countries, the infection rate was very high. In groups 2, 3, and 4 of countries, infection rates were high, medium, and low, respectively (Table 3). Ranking and grouping were performed at all six time points.

**Table 3.** Averages of ecological, economic, social characteristics in the 4 country-groups at 6 time points.

| Date | Country rank | Country group | Infection rate | Cases per 1 million people | GDP per capita \$ | Ecological Footprint gha/person | Biological Capacity gha/person | Urban population % | Median age years | Population density capita/km<sup>2</sup> |
| --- | --- | --- | --- | --- | --- | --- | --- | --- | --- | --- |
| 18-Apr-2020 | 1-35 | First | Very high | 1 567 | 38 871 | 5.6 | 3.1 | 76% | 40 | 172 |
| 2-May-2020 | 1-35 | First | Very high | 2 149 | 37 394 | 5.5 | 3.3 | 77% | 39 | 129 |
| 16-May-2020 | 1-35 | First | Very high | 2 797 | 37 610 | 5.4 | 3.2 | 78% | 39 | 169 |
| 30-May-2020 | 1-35 | First | Very high | 3 601 | 36 786 | 5.5 | 3.1 | 79% | 38 | 167 |
| 18-Jun-2020 | 1-35 | First | Very high | 4 943 | 33 254 | 5.2 | 3.7 | 78% | 37 | 129 |
| 4-Jul-2020 | 1-35 | First | Very high | 6 101 | 30 531 | 5.1 | 3.7 | 77% | 36 | 123 |
| 18-Apr-2020 | 36-70 | Second | High | 221 | 14 166 | 3.6 | 5.0 | 71% | 37 | 87 |
| 2-May-2020 | 36-70 | Second | High | 315 | 13 788 | 3.5 | 3.7 | 69% | 36 | 84 |
| 16-May-2020 | 36-70 | Second | High | 455 | 14 100 | 3.6 | 4.2 | 69% | 36 | 86 |
| 30-May-2020 | 36-70 | Second | High | 599 | 13 174 | 3.3 | 4.0 | 65% | 35 | 79 |
| 18-Jun-2020 | 36-70 | Second | High | 883 | 13 250 | 3.2 | 2.9 | 62% | 34 | 155 |
| 4-Jul-2020 | 36-70 | Second | High | 1 305 | 15 099 | 3.2 | 5.2 | 63% | 33 | 158 |
| 18-Apr-2020 | 71-105 | Third | Medium | 34 | 3 628 | 1.9 | 5.1 | 55% | 24 | 138 |
| 2-May-2020 | 71-105 | Third | Medium | 64 | 5 310 | 2.1 | 3.9 | 55% | 26 | 192 |
| 16-May-2020 | 71-105 | Third | Medium | 114 | 4 655 | 1.8 | 3.7 | 53% | 26 | 149 |
| 30-May-2020 | 71-105 | Third | Medium | 173 | 6 259 | 2.1 | 4.0 | 57% | 28 | 154 |
| 18-Jun-2020 | 71-105 | Third | Medium | 278 | 9 651 | 2.5 | 6.9 | 60% | 29 | 108 |
| 4-Jul-2020 | 71-105 | Third | Medium | 382 | 10 525 | 2.6 | 4.8 | 59% | 31 | 110 |
| 18-Apr-2020 | 106-140 | Fourth | Low | 4 | 1 922 | 1.7 | 2.3 | 40% | 22 | 96 |
| 2-May-2020 | 106-140 | Fourth | Low | 7 | 2 108 | 1.7 | 4.7 | 41% | 22 | 88 |
| 16-May-2020 | 106-140 | Fourth | Low | 16 | 2 186 | 1.7 | 4.4 | 41% | 22 | 90 |
| 30-May-2020 | 106-140 | Fourth | Low | 27 | 2 332 | 1.8 | 4.3 | 41% | 22 | 95 |
| 18-Jun-2020 | 106-140 | Fourth | Low | 46 | 2 394 | 1.8 | 1.9 | 43% | 23 | 102 |
| 4-Jul-2020 | 106-140 | Fourth | Low | 68 | 2 384 | 1.8 | 1.8 | 42% | 23 | 104 |

The dynamics and management of the epidemic varied from country to country, so the composition of the groups varied. The four groups of countries were characterized by averages that were also not constant over time. In the four groups of countries, not only the infection rate but also the ecological, economic and social characteristics differed significantly (Table 3).

The infection rate in group 1 is very high and ranges from 1,567 to 6,101 cases per million people. In groups 2, 3, and 4, the infection rates were 221–1305, 34–382, and 4–68 cases / million people. The range of group 1 is significantly higher than that of the other 3 groups. There is no overlap between groups 1 and 2.

The GDP ranges per capita differ significantly in the 4 groups, so they do not overlap in either case. In groups 1, 2, 3, and 4, per capita GDP was $ 30,531–38,871, 13,174–15,099, 3,628–10,525, and 1,922– 2,394. The higher the range of GDP per capita, the higher the infection rate. GDP per capita characterizes production. This means that higher infection rates have been reported in groups of countries with higher per capita GDP and higher production.

The state of nature, the state of the environment is characterized by Ecological Footprint and Biological Capacity. In groups 1, 2, 3 and 4 of the countries, the Ecological Footprint per capita is 5.1 - 5.6, 3.2 - 3.6, 1.8 - 2.6 and 1.7 - 1, 8 global hectares and the ranges do not overlap. The larger the range of the Ecological Footprint, the higher the range of the infection rate. The Ecological Footprint characterizes consumption. Higher infection rates have been reported in groups of countries with higher Ecological Footprints and higher consumption.

In the four groups of countries, the urban population declined systematically and ranged from 76-79%, 62-71%, 53-60%, and 40-43%. Similarly, the median age decreased in the four country groups and ranged from 36 to 40, 33 and 37, 24 and 31, and 22 to 23 years.

The ranking of 140 countries by infection rate resulted in four groups of countries with different ecological, economic, and social characteristics.

For this reason, we also wondered whether the relationship between infection rate and ecological, economic, and social characteristics was also different. The multivariate relationships are shown in Table 4. In the first group of countries, the very high infection rate can be explained mainly by GDP per capita and the Ecological Footprint. In groups 2, 3, and 4, infection rates were mainly associated with social characteristics (median age and urban population). Table 4 does not show causal relationships, but confirms that the infection rate is not independent of ecological, economic, and social conditions.

**Table 4.** Multivariate regression equations between infection rate and ecological, economic, social characteristics in the 4 country-groups at 6 time points.

| Date | Country group | Infection rate | Regression equation | R<sup>2</sup> | F value | P value |
| --- | --- | --- | --- | --- | --- | --- |
| 18-Apr-2020 | First | Very high | Cases/1M = 0.0305*GDP/capita - 121.5*Biocapacity + 19.72*Median age | 0.8428 | 57 | 5.91E-13 |
| 2-May-2020 | First | Very high | Cases/1M = 0.0340*GDP/capita + 35.34*Median age - 152.8*Biocapacity | 0.8586 | 65 | 1.09E-13 |
| 16-May-2020 | First | Very high | Cases/1M = 490.8*Ecofootprint | 0.8085 | 144 | 9.43E-14 |
| 30-May-2020 | First | Very high | Cases/1M = 647.2*Ecofootprint | 0.7340 | 94 | 2.62E-11 |
| 18-Jun-2020 | First | Very high | Cases/1M = 1471*Ecofootprint - 0.0843*GDP/capita | 0.7041 | 39 | 1.87E-09 |
| 4-Jul-2020 | First | Very high | Cases/1M = 1744*Ecofootprint - 0.0990*GDP/capita | 0.6930 | 37 | 3.45E-09 |
| 18-Apr-2020 | Second | High | Cases/1M = 5.957*Median age | 0.7683 | 113 | 2.46E-12 |
| 2-May-2020 | Second | High | Cases/1M = 8.817*Median age | 0.7887 | 127 | 5.07E-13 |
| 16-May-2020 | Second | High | Cases/1M = 12.45*Median age | 0.7935 | 131 | 3.42E-13 |
| 30-May-2020 | Second | High | Cases/1M = 14.26*Median age + 21.39*Biocapacity | 0.8454 | 90 | 4.18E-14 |
| 18-Jun-2020 | Second | High | Cases/1M = 1386*Urban pop. | 0.8933 | 285 | 4.30E-18 |
| 4-Jul-2020 | Second | High | Cases/1M = 2005*Urban pop. | 0.8956 | 292 | 2.99E-18 |
| 18-Apr-2020 | Third | Medium | Cases/1M = 34.35*Urban pop. + 8.940*Ecofootprint - 0.3819*Biocapacity | 0.9232 | 128 | 6.45E-18 |
| 2-May-2020 | Third | Medium | Cases/1M = 1.190*Median age + 53.08*Urban pop. + 0.8396*Biocapacity | 0.9066 | 104 | 1.48E-16 |
| 16-May-2020 | Third | Medium | Cases/1M = 4.076*Median age | 0.8319 | 168 | 1.01E-14 |
| 30-May-2020 | Third | Medium | Cases/1M = 5.867*Median age | 0.8432 | 183 | 3.08E-15 |
| 18-Jun-2020 | Third | Medium | Cases/1M = 8.649*Median age | 0.8062 | 141 | 1.16E-13 |
| 4-Jul-2020 | Third | Medium | Cases/1M = 11.34*Median age | 0.7774 | 119 | 1.24E-12 |
| 18-Apr-2020 | Fourth | Low | Cases/1M = 1.478*Ecofootprint + 0.0141*Pop.density | 0.7164 | 42 | 9.31E-10 |
| 2-May-2020 | Fourth | Low | Cases/1M = 13.65*Urban pop. + 0.0184*Pop.density | 0.6812 | 35 | 6.43E-09 |
| 16-May-2020 | Fourth | Low | Cases/1M = 6.038*Ecofootprint + 0.0372*Pop.density | 0.5750 | 22 | 7.40E-07 |
| 30-May-2020 | Fourth | Low | Cases/1M = 59.03*Urban pop. | 0.6166 | 55 | 1.42E-08 |
| 18-Jun-2020 | Fourth | Low | Cases/1M = 99.73*Urban pop. | 0.7093 | 83 | 1.21E-10 |
| 4-Jul-2020 | Fourth | Low | Cases/1M = 149.4*Urban pop. | 0.7403 | 97 | 1.73E-11 |

When the global world was analyzed as a single ecosystem, the regression equations were able to explain 50 - 70% of the infection rate (Table 2). However, categorization of countries by infection rate resulted in a closer correlation with R^2^ growth (Table 4). In the four groups of countries, the selected variables explain 69–86%, 76–90%, 77–93%, and 57–75% of the infection rate. The regression equations of 4 groups of countries better describe the relationship between infection rate and ecological, economic, social characteristics.

### Country-group with very high infection rate

We found that the four groups of countries have different ecological, economic, and social characteristics. However, the first group deserves special attention due to the very high infection rate as well as the highest per capita GDP and Ecological Footprint (Table 3). This group is dominated by developed countries with the highest production (GDP/capita) and consumption (Ecological Footprint/person). But why was the infection rate highest in the developed group of countries?

High production and high consumption obviously cannot cause infection. In developed countries, however, high industrial and agricultural production and high consumption have long had a direct and indirect impact on nature. In other words, nature has long been under great pressure in developed countries. As a result of the cumulative effects, the ecological characteristics of the ecosystem may have changed. This is supported by the data in Table 3. The Ecological Footprint is significantly higher in the first group of countries (5.6 - 5.1) than in the other three groups of countries (3.6 - 1.7). In addition, the ecosystem is unbalanced because the Ecological Footprint is always much higher than the Biological Capacity. This is not the case in the other three groups of countries, where the Ecological Footprint is in most cases lower than the Biological Capacity. So, the importance of ecological characteristics is not negligible, especially in developed countries.

Therefore, it was reasonable to study the ecological properties of the 1^st^ group of countries in more detail. We can analyze the ecological characteristics of a total of 46 countries (Table 5). Countries are ranked according to infection rates. The Ecological Footprint exceeds 4 in 32 countries. The Ecological Footprint is highest (> 8) in Luxemburg, Qatar, United Arab Emirates and United States. Biological Capacity < 1 in 10 countries where the ecosystem is not able to produce enough biological material for the population. A country’s ecosystem is unbalanced if the Ecological Footprint (i.e., human consumption) is much larger than its Biological Capacity. This occurs in 11 countries where the infection rate is also very high. Biocapacity shortages, i.e., overconsumption, are very high (< −4) in Luxembourg, Qatar, the United Arab Emirates, Belgium, the United Kingdom, the United States, Israel, the Netherlands and Saudi Arabia, Oman and Malta. Excessive consumption in these countries results in ecosystem imbalances.

**Table 5.** Ecological characteristics of countries reporting very high infection rates (6 time points).

| **18-Apr-2020** | Cases | Ecological | Biological | **2-May-2020** | Cases | Ecological | Biological | **16-May-2020** | Cases | Ecological | Biological |
| --- | --- | --- | --- | --- | --- | --- | --- | --- | --- | --- | --- |
|  | per 1 million | Footprint | Capacity |  | per 1 million | Footprint | Capacity |  | per 1 million | Footprint | Capacity |
|  | people | gha/person | gha/person |  | people | gha/person | gha/person |  | people | gha/person | gha/person |
| Luxembourg | 5 650 | 15.3 | 1.8 | Luxembourg | 6 074 | 15.3 | 1.8 | Qatar | 10 774 | 14.4 | 1.0 |
| Spain | 4 158 | 4.0 | 1.4 | Spain | 5 252 | 4.0 | 1.4 | Luxembourg | 6 280 | 15.3 | 1.8 |
| Belgium | 3 322 | 6.3 | 0.8 | Qatar | 5 162 | 14.4 | 1.0 | Spain | 5 914 | 4.0 | 1.4 |
| Switzerland | 3 166 | 4.6 | 1.0 | Belgium | 4 273 | 6.3 | 0.8 | Ireland | 4 859 | 5.1 | 3.4 |
| Ireland | 2 989 | 5.1 | 3.4 | Ireland | 4 219 | 5.1 | 3.4 | Belgium | 4 747 | 6.3 | 0.8 |
| Italy | 2 910 | 4.4 | 0.9 | Switzerland | 3 445 | 4.6 | 1.0 | USA | 4 498 | 8.1 | 3.6 |
| France | 2 325 | 4.5 | 2.4 | Italy | 3 431 | 4.4 | 0.9 | Italy | 3 702 | 4.4 | 0.9 |
| USA | 2 232 | 8.1 | 3.6 | USA | 3 420 | 8.1 | 3.6 | UK | 3 540 | 6.6 | 1.1 |
| Portugal | 1 931 | 4.1 | 1.3 | UK | 2 614 | 6.6 | 1.1 | Switzerland | 3 536 | 4.6 | 1.0 |
| Netherlands | 1 844 | 4.9 | 0.8 | France | 2 564 | 4.5 | 2.4 | Belarus | 3 035 | 4.6 | 3.4 |
| Qatar | 1 738 | 14.4 | 1.0 | Portugal | 2 486 | 4.1 | 1.3 | Sweden | 2 941 | 6.4 | 9.6 |
| Germany | 1 715 | 4.9 | 1.6 | Netherlands | 2 348 | 4.9 | 0.8 | Portugal | 2 824 | 4.1 | 1.3 |
| UK | 1 682 | 6.6 | 1.1 | Sweden | 2 186 | 6.4 | 9.6 | France | 2 751 | 4.5 | 2.4 |
| Israel | 1 544 | 4.9 | 0.3 | Germany | 1 958 | 4.9 | 1.6 | Netherlands | 2 561 | 4.9 | 0.8 |
| Sweden | 1 369 | 6.4 | 9.6 | Israel | 1 866 | 4.9 | 0.3 | UAE | 2 211 | 8.9 | 0.6 |
| Panama | 990 | 2.3 | 2.8 | Belarus | 1 675 | 4.6 | 3.4 | Panama | 2 152 | 2.3 | 2.8 |
| Turkey | 976 | 3.4 | 1.5 | Panama | 1 557 | 2.3 | 2.8 | Germany | 2 099 | 4.9 | 1.6 |
| Canada | 885 | 7.7 | 15.1 | Ecuador | 1 493 | 1.8 | 2.0 | Chile | 2 071 | 4.3 | 3.5 |
| UAE | 686 | 8.9 | 0.6 | Canada | 1 459 | 7.7 | 15.1 | Canada | 1 979 | 7.7 | 15.1 |
| Moldova | 589 | 1.7 | 1.2 | Turkey | 1 451 | 3.4 | 1.5 | Israel | 1 922 | 4.9 | 0.3 |
| Ecuador | 511 | 1.8 | 2.0 | UAE | 1 318 | 8.9 | 0.6 | Ecuador | 1 787 | 1.8 | 2.0 |
| Chile | 509 | 4.3 | 3.5 | Moldova | 987 | 1.7 | 1.2 | Turkey | 1 739 | 3.4 | 1.5 |
| Belarus | 506 | 4.6 | 3.4 | Chile | 890 | 4.3 | 3.5 | Moldova | 1 424 | 1.7 | 1.2 |
| **30-May-2020** | Cases | Ecological | Biological | **18-Jun-2020** | Cases | Ecological | Biological | **4-Jul-2020** | Cases | Ecological | Biological |
|  | per 1 million | Footprint | Capacity |  | per 1 million | Footprint | Capacity |  | per 1 million | Footprint | Capacity |
|  | people | gha/person | gha/person |  | people | gha/person | gha/person |  | people | gha/person | gha/person |
| Qatar | 19 211 | 14.4 | 1.0 | Qatar | 30 074 | 14.4 | 1.0 | Qatar | 35 324 | 14.4 | 1.0 |
| Luxembourg | 6 425 | 15.3 | 1.8 | Chile | 11 779 | 4.3 | 3.5 | Chile | 15 070 | 4.3 | 3.5 |
| Spain | 6 124 | 4.0 | 1.4 | USA | 6 791 | 8.1 | 3.6 | USA | 8 833 | 8.1 | 3.6 |
| USA | 5 459 | 8.1 | 3.6 | Luxembourg | 6 540 | 15.3 | 1.8 | Panama | 8 342 | 2.3 | 2.8 |
| Ireland | 5 054 | 5.1 | 3.4 | Spain | 6 253 | 4.0 | 1.4 | Luxembourg | 7 150 | 15.3 | 1.8 |
| Belgium | 5 022 | 6.3 | 0.8 | Belarus | 5 996 | 4.6 | 3.4 | Sweden | 7 071 | 6.4 | 9.6 |
| Chile | 4 966 | 4.3 | 3.5 | Sweden | 5 550 | 6.4 | 9.6 | Belarus | 6 696 | 4.6 | 3.4 |
| Belarus | 4 408 | 4.6 | 3.4 | Panama | 5 240 | 2.3 | 2.8 | Spain | 6 366 | 4.0 | 1.4 |
| UK | 4 021 | 6.6 | 1.1 | Belgium | 5 208 | 6.3 | 0.8 | Belgium | 5 335 | 6.3 | 0.8 |
| Italy | 3 848 | 4.4 | 0.9 | Ireland | 5 137 | 5.1 | 3.4 | Ireland | 5 166 | 5.1 | 3.4 |
| Sweden | 3 677 | 6.4 | 9.6 | UK | 4 427 | 6.6 | 1.1 | UAE | 5 142 | 8.9 | 0.6 |
| Switzerland | 3 566 | 4.6 | 1.0 | UAE | 4 426 | 8.9 | 0.6 | Moldova | 4 381 | 1.7 | 1.2 |
| UAE | 3 431 | 8.9 | 0.6 | Italy | 3 939 | 4.4 | 0.9 | Portugal | 4 273 | 4.1 | 1.3 |
| Portugal | 3 157 | 4.1 | 1.3 | Portugal | 3 735 | 4.1 | 1.3 | UK | 4 197 | 6.6 | 1.1 |
| Panama | 2 908 | 2.3 | 2.8 | Switzerland | 3 606 | 4.6 | 1.0 | Italy | 3 993 | 4.4 | 0.9 |
| France | 2 863 | 4.5 | 2.4 | Moldova | 3 249 | 1.7 | 1.2 | Switzerland | 3 720 | 4.6 | 1.0 |
| Netherlands | 2 700 | 4.9 | 0.8 | Netherlands | 2 878 | 4.9 | 0.8 | Ecuador | 3 438 | 1.8 | 2.0 |
| Canada | 2 391 | 7.7 | 15.1 | Ecuador | 2 785 | 1.8 | 2.0 | Israel | 3 156 | 4.9 | 0.3 |
| Ecuador | 2 189 | 1.8 | 2.0 | Canada | 2 654 | 7.7 | 15.1 | Netherlands | 2 938 | 4.9 | 0.8 |
| Germany | 2 187 | 4.9 | 1.6 | France | 2 431 | 4.5 | 2.4 | Canada | 2 787 | 7.7 | 15.1 |
| Moldova | 2 007 | 1.7 | 1.2 | Germany | 2 263 | 4.9 | 1.6 | France | 2 558 | 4.5 | 2.4 |
| Turkey | 1 936 | 3.4 | 1.5 | Turkey | 2 183 | 3.4 | 1.5 | Turkey | 2 426 | 3.4 | 1.5 |
| Israel | 1 850 | 4.9 | 0.3 | Israel | 2 163 | 4.9 | 0.3 | Germany | 2 354 | 4.9 | 1.6 |

In 23 countries, the infection rate was very high at all 6 time points. Of these 23 countries, 14 belong to Europe, 4 to Asia (Middle East), 3 to North America and 2 to South America. This group of countries is dominated by developed countries and their ecological characteristics have proven to be particularly significant. In countries where the infection rate is very high, the Ecological Footprint is high, or the Biological Capacity is low, or the ecosystem is unbalanced.

Table 5 is dominated by developed countries. The gross domestic product of the United States, Germany, the United Kingdom, France and Italy is high and among the top ten in the world. Their estimated and reported infection rates as a function of time are compared in Figure 1. For the estimation, we used the regression equations of the first group of countries (Table 4) as well as the ecological, economic and social characteristics of the given country.

**Figure 1.**
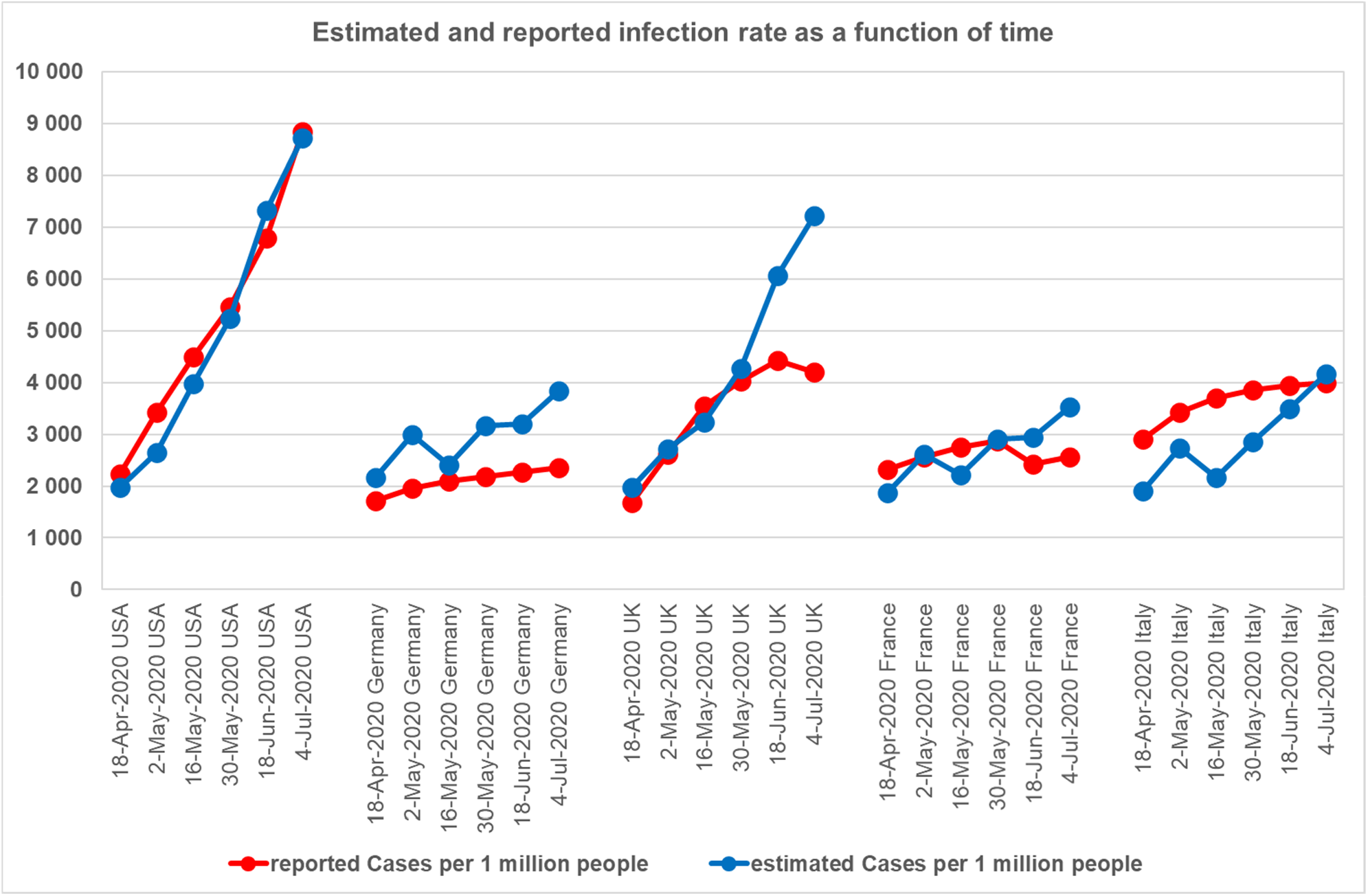
Reported and estimated infection rates at six time points. Legend: X-axis: country and six time points. Y-axis: reported and estimated infection rate (cases per one million people).

The United States is the strongest economy in the world. The reported rate of infections per million people increased from 2,232 to 8,833, covering a wide range. The estimated and reported infection rates fit well. This indicates that the rate of infection in the world’s strongest economy is not independent of ecological, economic, and social characteristics. In Germany, the reported infection rate is increasing slowly and gradually, from 1,715 to 2,354 cases / million people. Estimated infections resulted in higher than reported infections. These may indicate effective management of the epidemic. In the UK, the reported infection rate initially increased rapidly and the estimated and reported infections correlated well. It is likely that the appropriate restrictions then stabilized the rate of infection. In France, the number of reported infections was less than 2,600 cases per million people. Estimated and reported infections were twice correlated, twice underestimated, and twice overestimated. In Italy, the infection rate was high on 18 April 2020 (2,910 cases per million people were reported) and the reported infection was higher than the estimated infection. Subsequently, management of the epidemic gradually stabilized the rate of infection, and reported and estimated infections approached each other.

To the best of our knowledge, the relationship between COVID-19 pandemic and ecological, economic, and social characteristics has not yet been studied. In this case, however, our concept and results need to be interpreted and discussed from a scientific perspective.

## Discussion

### Sustainable ecosystems

We interpret the world as a single global ecosystem made up of ecosystem units representing countries. But the global ecosystem is under threat because human activities are affecting the Earth. Steffen et al concluded that the boundaries of our planet are levels of man-made perturbations beyond which the functioning of the Earth can change significantly [5]. According to their calculations, changes in climate and biosphere integrity are at high risk, while changes in biogeochemical flows and the terrestrial system pose an increased threat to Earth’s stability. In 2020, the Intergovernmental Science-Policy Platform on Biodiversity and Ecosystem Services (IPBES) proposed a reassessment of the relationship between man and nature, as ecological disturbances and unsustainable consumption result in biodiversity loss, climate change and pandemic risk [6]. Countries’ ecosystems are also at risk, especially in developed countries, where people have a very significant impact on the natural ecosystem, its natural components, the atmosphere, water, soil and living things. Unfortunately, adverse effects are usually recognized late (acidification, air, water, soil pollution, etc.) [7, 8].

These adverse effects have ecological consequences because nature is a sustainable, logically functioning ecosystem and responds to external influences. The resistance of natural components to adverse effects follows the order of buffer capacity: air < water < soil < living organism. Air is the most sensitive to pollution because its buffer capacity is low. However, these adverse effects have not only ecological but also economic consequences. In the U.S. economy, for example, air pollution has become less intense. As a result, Gross Economic Damage (due to premature death) decreased [9]. That is, air pollution has caused economic losses. The ecological consequences therefore had direct economic consequences. The economic consequences of COVID-19 can be assessed in light of the above. The pandemic can be understood as a reaction of nature. Based on this, the loss of GDP caused by COVID-19 pandemic can be interpreted as a tax reimbursed to nature.

Sustainable natural ecosystems (terrestrial ecosystems, aquatic ecosystems) are based on the ecological pyramid formed during evolution [10], where production and consumption are in balance. But both evolution and the ecological pyramid are built on the invisible world of microorganisms. As a result, soil, plant, animal, and human life are inseparable from the microscopic world. Microorganisms (as well as viruses, including pathogens) live in the same environment as us.

However, urban ecosystems have been created by humans [11]. The urban population lives in the urban ecosystem. It is worth noting that the characteristics of urban ecosystems and the first group of countries are similar: per capita GDP (economic production) is high, Ecological Footprint (human consumption) is high, Biological Capacity is low, and the ecosystem is unbalanced. This may suggest an indirect relationship between infection rate and ecological characteristics of the urban ecosystem, but this indirect relationship cannot be studied with our available data.

In the urban ecosystem, regardless of ecological characteristics, infection can spread much more easily due to high population density, small social distance, and large number of human contacts. These direct reasons explain why the rate of COVID-19 coronavirus infection may be related to the urban population.

In our analysis, the urban population represents the urban ecosystem. Our regression equations in Table 4 often include the urban population. For example, in group 4 of countries, the urban population is included four times in the regression equation. In these 35 countries, the urban population is less than 43%, so the majority of people live in rural areas where the infection cannot spread easily, so the infection rate is also low. Our results suggest that the role of the urban population in the epidemic was significant, especially in countries with low urbanization.

Although the Ecological Footprint and Biological Capacity of urban ecosystems are unknown, their ecological characteristics deserve increased attention. Flies et al. [12] showed that urbanization reduces the abundance and diversity of airborne microbes and suggested studying the impact of aerobiome on human health in urban ecosystems. Robinson et al. [13] studied the aerobiome of urban green spaces and found significant vertical stratification in the potentially pathogenic and beneficial bacterial taxa.

### Microorganisms in ecosystems and humans

Sustainable natural ecosystems, their ecological pyramids and their evolution are based on microorganisms. Microorganisms play a vital role in both ecosystems and humans. In 2020, the definition of microbiome has been updated. A microbiome represents a microbiota (community of microorganisms) as well as an “activity theater” that also contains microbial structural elements (including mobile genetic elements such as viruses), microbial metabolites, and environmental conditions [14]. The understanding of microbiomes in soils, plants, animals and humans might play a key role in solving new challenges, which are associated with anthropogenic driven changes in the field of planetary health. In this way, nature’s resilience to adverse effects depends on the function, sensitivity, and stability of the ecosystem’s microbiome as well. So, microorganisms play a significant role in the global ecosystem. For example, microbial motors drive the biogeochemical cycles of the Earth [15] and microbial activity mediates the fluxes of greenhouse gases [16]. Recognizing the global role of microorganisms, we direct our attention to the interaction between microorganisms and the components of nature.

**Coexisting microorganisms in the soil** form the basis of the life of healthy terrestrial ecosystems [10, 17]. Soil life is the fundamental of the ecological pyramid of terrestrial ecosystems. In stressed ecosystems the soil’s ability to recover is the key to maintain soil health [18]. Billions of micro-organisms in soil form a complex community in which hundreds of microbial species coexist. The community of soil microorganisms (soil microbiota) lives in harmony with its surrounding environment. The long-term quality of soil is determined by its physical, chemical and biological properties [19]. Soil habitats probably contain the greatest microbial diversity on Earth [20]. Soil archaea, bacteria, fungi, and viruses are globally as well as locally diverse [21]. Harmful impacts on soil microbiota affect the biodiversity of soil flora and fauna, as related to soil and plant health [22]. Soil fertility is greatly determined by the microbiological activity of the root environment in both healthy [23] and in polluted soil [24]. Microbial diversity is critical for maintaining the multifunctionality of terrestrial ecosystems [25]. Although soil vitality is dominated by microorganisms, their coexistence with soil fauna is not negligible either.

**Coexisting microorganisms with animals** influence animal health. Biodiversity in agricultural landscapes affects the protective microbiome of insects [26]. The insect – microbiome interaction influences the host and the microbial symbiont [27]. The microbiome associated with insects is very complex and behaves as a mini ecosystem [28], that is highly dependent on the environment [29]. The biodiversity of the host-associated microbiota is recognized as an essential component of wildlife management, that has a profound impact on animal health, but these microbial communities can be drastically altered by anthropogenic activities [30]. Unfortunately, the reality is that insects have declined by 40% in recent decades, and one-third are at risk [31]. Global threats to insects are caused by a number of factors (agricultural intensification, insecticides, pollution, deforestation, urbanization, etc.) [32]. Pesticide exposure, infectious disease, and nutritional stress contribute to honey bee mortality and a high rate of colony loss. Symbionts may be major regulators of stress tolerance and disease resistance. Missing microbes in bees, as well as systematic depletion of key symbionts impair bee immunity. Treatment strategies based on microbiota restoration are promising in restoring bee colony health [33]. Healthy microbiomes represent the foundations of soil, plant and animal life in sustainable ecosystems. However, this is true not only for the components of the ecosystem, but also for healthy people.

**Coexisting microbes with humans** support human health. Both people and society are seen as part of the country’s ecosystem unit. Human and ecosystem health are not independent of each other. Relman [34] adopted ecological perspectives to understand the health functions of the human microbiota and the resilience of the human microbial ecosystem. The number of microbes (which live inside and outside our body) is about ten times greater than the number of our body cells [35]. Therefore, the human microbiome has become the subject of intensive research to clarify their role in health and disease. For example, the skin acts as a physical barrier, preventing the invasion of foreign pathogens while providing a home to the commensal microbiota [36]. Pulmonary immunity is shaped by interaction with the microbiota [37]. Communication disorders between the innate immune system and the intestinal microbiota may contribute to the development of complex diseases [38].

The microbiota plays an essential role in the functioning of the host immune system, not only in case of animals but also in case of humans. However, in high-income countries, overuse of antibiotics, dietary changes, and so on have resulted in the selection of a microbiota that lacks the resilience and diversity needed to establish balanced immune responses [39]. Our diet influences the gut microbiome and the immune system [40]. The Western diet activates the innate immune system and impairs adaptive immunity, leading to chronic inflammation and impaired defense against viruses. Wider access to healthy food should be a top priority [41]. (However, healthy plants can be grown in healthy soil that is full of life.) Age-related changes in the intestinal microbiota were associated with the immune system in old age [42]. The immune system deteriorates with age and causes about 90% of excessive deaths in people over the age of 65 during a regular influenza season [43].

Our results are consistent with the scientific literature, because COVID-19 coronavirus infection can often be explained by median age (Table 4). For example, in groups 3 and 4, the median age was less than 31 years and the infection rate was medium or low (Table 3). The younger the population, the stronger the immune system and the lower the infection rate. Assuming that humans are seen as part of the natural ecosystem, it is not surprising that the resilience of both natural ecosystems and humans is closely related to the world of a healthy microbial community.

### The future

The COVID-19 coronavirus pandemic is a serious warning to humanity, who can choose from three main directions of development. The world of the future can be economy-driven (production-based) or society-driven (consumption-based) or nature-driven (coexistence-based). This can be deduced from our results and the scientific literature. Because COVID-19 infection is very high in developed countries, the coronavirus epidemic indicates to humanity that the only chance of survival is to live in a coexisting world. Both the economy and society must be in harmony with nature, creating sustainable ecosystems in developed countries as well.

This makes not only scientific but also economic sense. In 2020, the World Economic Forum assessed global risks in terms of probability and impact. It has been recognized that risks related to nature are underestimated in business decision making and the new nature economy needs to take into account the economic value of nature. According to the World Economic Forum, business rationality lies in the preservation or restoration of natural ecosystems [31].

## Data Availability

All data are publicly available.

https://www.worldometers.info/coronavirus/

## Notes

### Competing Interest Statement

The authors have declared no competing interest.

### Funding Statement

The research was not funded.

